# Assessing the feasibility of sustaining SARS-CoV-2 local containment in China in the era of highly transmissible variants

**DOI:** 10.1101/2022.05.07.22274792

**Authors:** Yan Wang, Kaiyuan Sun, Zhaomin Feng, Lan Yi, Yanpeng Wu, Hengcong Liu, Quanyi Wang, Marco Ajelli, Cécile Viboud, Hongjie Yu

## Abstract

We developed a spatially structured, fully stochastic, individual-based SARS-CoV-2 transmission model to evaluate the feasibility of sustaining SARS-CoV-2 local containment in mainland China considering currently dominant Omicron variants, China’s current immunization level, and non-pharmaceutical interventions (NPIs). We also built a statistical model to estimate the overall disease burden under various hypothetical mitigation scenarios. We found that due to high transmissibility, neither Omicron BA.1 or BA.2 could be contained by China’s pre-Omicron NPI strategies which were successful prior to the emergence of the Omicron variants. However, increased intervention intensity, such as enhanced population mobility restrictions and multi-round mass testing, could lead to containment success. We estimated that an acute Omicron epidemic wave in mainland China would result in significant number of deaths if China were to reopen under current vaccine coverage with no antiviral uptake, while increasing vaccination coverage and antiviral uptake could substantially reduce the disease burden. As China’s current vaccination has yet to reach high coverage in older populations, NPIs remain essential tools to maintain low levels of infection while building up protective population immunity, ensuring a smooth transition out of the pandemic phase while minimizing the overall disease burden.

---

Prior to the emergence of the SARS-CoV-2 Omicron variants, a few countries and regions had successfully maintained local containment with effective border controls and non-pharmaceutical interventions (NPIs), while the rest of the world adopted mitigation approaches that resulted in higher burden of disease and loss of lives ^1^. The state of the pandemic since late 2021 has posed more challenges on the long-term sustainability of SASR-CoV-2 local containment: the emergence and rapid spread of the highly transmissive and immune evasive Omicron variants, together with the observed waning of vaccine-induced immunity, have demonstrated that none of the currently licensed vaccines can guarantee population immunity levels capable of preventing major epidemic waves, even with high booster coverage rates ^2,3^. Furthermore, reports of spillover of human SARS-CoV-2 viruses into a wide range of animal hosts, followed by sustained transmission in animals, as well as reverse zoonosis from animals to humans, highlight the existence of multiple animal reservoirs for SARS-CoV-2 ^4-7^. All the available evidence points to the inevitable long-term circulation of SARS-CoV-2 in human populations.

A decoupling of immune protection against infection from immune protection against severe outcomes has also been observed, suggesting that while an “immunity wall” against SARS-CoV-2 infection is likely unattainable, an “immunity wall” against severe outcomes is still achievable through vaccination with booster shots ^2,8^, or through natural immunity for countries where vaccine coverage has remained low so far. In fact, countries such as Australia, New Zealand, and Singapore achieved high immunization coverage prior to a successful transition from local containment to mitigation strategies, reporting remarkably low number of deaths despite a large number of SARS-CoV-2 infections ^9^. In contrast, the fifth COVID-19 wave in Hong Kong Special Administrative Region (Hong Kong SAR), China, caused by the Omicron variants, overcame the long-standing success of keeping the majority of the population free of SARS-CoV-2 infections (Supplementary Fig. 1a). A particularly low vaccine coverage among the elderly has resulted in high rates of severe COVID-19 disease, overwhelming the healthcare system and causing a considerable number of deaths ^10^.

Until mid-March 2022, mainland China successfully sustained local containment since the initial Wuhan epidemic wave through rigorous border entry quarantine/isolation and swift suppression of sporadic local outbreaks, including those caused by both Delta and Omicron variants. However, a major Omicron epidemic started in late-February 2022, leading to over 620,000 reported infections (as of May 16, 2022) in Shanghai ^11^, causing the government to put the entire city into lockdown for more than a month. Moreover, similar to Hong Kong, substantial immunity gaps exist in mainland China, especially in the most vulnerable populations (the elderly and/or individuals with underlying medical conditions). In the population over the age of 80 years, only 50.7% have received primary shots of inactivated vaccines; only 19.7% had received a booster shot as of March 17, 2022 (Supplementary Fig. 2); note that due to initial immunization prioritization to healthy adults, vaccine coverage declines with age in China ^12^. A rushed reopening of international borders and lifting of existing NPIs could potentially lead to catastrophic consequences in mainland China, similar to those observed during the fifth wave in Hong Kong, but on a far larger scale: over 1/6 of the world population lives in mainland China. Whether it is possible for China to sustain a local containment policy in face of the highly transmissible and/or immune evasive Omicron variants, and under what conditions, is of paramount importance to resolve at this stage of the pandemic. The ability to prevent major epidemic waves until a safe transition to mitigation strategies can be safely ensured is essential to minimize the overall disease burden and societal cost of COVID-19 in China.

## Results

In this study, we utilized a previously developed, spatially structured, fully stochastic, individual-based SARS-CoV-2 transmission model that was used to reconstruct the containment effort of the Xinfadi outbreak in Beijing, China, caused by the ancestral lineage in a population with negligible immunity ^13^. We updated the model to consider the increased transmissibility and immune evasion of Omicron’s BA.1 and BA.2 sublineages, as well as vaccination effectiveness and coverage representative of current and future immunization efforts in China. We further created five levels of NPIs with incrementally increasing intensity from a baseline corresponding to the levels adopted during the Xinfadi outbreak. The findings of this work can be generalized to anticipate the counter-measure requirements necessary to contain future outbreaks in densely populated urban settings experiencing variants that have characteristics similar to Omicron. (detailed in Methods Section 1).

Specifically, we examine the feasibility of achieving outbreak containment conditional on SARS-CoV-2 variant introduction in Beijing, a highly urbanized setting, under various scenarios. We considered two SARS-CoV-2 variant scenarios: 1) the BA.1 sublineage of Omicron (*R*_0_ = 7.5, Methods Section 1.5), 2) the BA.2 sublineage of Omicron (*R*_0_ = 9.5, Methods Section 1.5). For each variant scenario, we further explore the feasibility of outbreak control under different NPI intensities and vaccine coverage. For NPIs, we start with the baseline levels that were implemented in 2020 to control the spread of the ancestral SARS-CoV-2 lineages, at a time where population immunity was minimal ^13^. We then strengthened NPIs gradually by improving: 1) symptom surveillance, 2) mask wearing, 3) occupation-based screening, 4) mass testing, and 5) mobility restrictions; these represent ascending NPI intensities from Level 1 through 5 (Table 1). Finally, for each NPI scenario we further explored a baseline immunization scenario with 100% population coverage of the primary vaccine course and two enhanced immunization scenarios with 100% population coverage through a booster dose of the Chinese-manufactured inactivated vaccines (homologous booster) or a booster dose of a viral vector vaccine (heterologous booster). We used estimates published in the study by Cai et. al. ^14^ for vaccine effectiveness against infection (*VE*_*I*_), symptomatic disease (*VE*_*s*_) and onward transmission (*VE*_*T*_) caused by Omicron variants. We assumed *VE*_*T*_=0%, *VE*_*I*_=9.1% and *VE*_*s*_=17.3% in our baseline immunization scenario, *VE*_*T*_=12.4%, *VE*_*I*_=17% and *VE*_*s*_=30.8% in the homologous booster scenario, and *VE*_*T*_=12.4%, *VE*_*I*_=56.1% and *VE*_*s*_=60.7% in the heterologous booster scenario (Supplementary Table 1). The detailed description of the analyzed scenarios is reported in the Methods Section 1.7. We use the effective reproduction number *R*_*eff*_ (defined as the average of the reproduction number for each individual infected after the implementation of NPIs) as the metric to determine the success of SARS-CoV-2 containment for each variant, NPI intensity, and immunization scenario, with *R*_*eff*_ < 1 indicating successful containment. To characterize the stochastic nature of SARS-CoV-2 transmission, we simulated each scenario 100 times and recorded the probability of successful containment.

**Table 1.** Intervention parameters of each scenario.

| Non-pharmaceutical intervention | Level 0 | Level 1 | Level 2 | Level 3 | Level 4 | Level 5 |
| --- | --- | --- | --- | --- | --- | --- |
| **Symptom surveillance** |  |  |  |  |  |  |
| Fraction of detected symptomatic infections | 33.3% | 66.7% | 66.7% | 66.7% | 66.7% | 66.7% |
| Mean time delay from symptom onset to hospitalization (days) | 3.7<sup>†</sup> | 2.7<sup>‡</sup> | 2.7 | 2.7 | 2.7 | 2.7 |
| **Mask wearing (% of population)** |  |  |  |  |  |  |
| In the workplace | 10% | 10% | 50% | 50% | 50% | 50% |
| In the community | 30% | 30% | 80% | 80% | 80% | 80% |
| **Occupational screening (% of working-age populations)** |  |  |  |  |  |  |
| High risk | 2.5% | 2.5% | 2.5% | 5% | 5% | 5% |
| Medium risk | 7.5% | 7.5% | 7.5% | 20% | 20% | 20% |
| **Mass screening** |  |  |  |  |  |  |
| Rounds | 1 | 1 | 1 | 1 | 5 | 5 |
| Geographical range | Street/township | Street/township | Street/township | Street/township | District/county | District/county |
| **Mobility restrictions<sup>#</sup>** |  |  |  |  |  |  |
| High-risk region | High | High | High | High | High | Lockdown |
| Moderate-risk region | Moderate | Moderate | Moderate | Moderate | Moderate | High |
| Low-risk region | Low<sup>*</sup> | Low | Low | Low | Low | Moderate |

Note: <sup>†</sup> The time delay from symptom onset to hospitalization is taken from a Weibull distribution (shape=2.38, scale=4.17); <sup>‡</sup> The time delay from symptom onset to hospitalization is taken from a gamma distribution (shape=0.69,rate=0.26); <sup>#</sup> The hypothetical origin-destination mobility matrices of NPI intensity Level 0-4 and Level 5 are shown in Supplementary Table 3 and Supplementary Table 4, respectively; <sup>\*</sup> We assume a 20% reduction in human mobility between low risk areas after the official report of the first infection according to the mobile phone signaling data.

In Fig. 1, we report the estimated probability of outbreak containment for each of the analyzed scenarios. Under the baseline immunization scenario, we found that both Omicron BA.1 and BA.2 could not be contained under the baseline NPI intensity (Fig. 1a, Level 0). Layering enhanced symptom surveillance, mask wearing mandate, and high-risk occupation screening (corresponding to Level 3 NPI intensity, Table 1) does not lead to significant improvement, with the probability of containment remaining negligible (3% and 1% for Omicron BA.1 and BA.2 outbreak, respectively). Multiple rounds of mass testing with an expanded geographical scope (corresponding to Level 4 NPI intensity, Table 1), would substantially increase the containment probability of Omicron BA.1 (36%), while the probability of containing Omicron BA.2 outbreak is still low (1%). Implementation of more stringent population mobility restrictions would increase the likelihood of containment to 91% and 82% for BA.1 and BA.2 respectively (Fig. 1a, Level 5). As of April 27, 2022, there are two major Omicron BA.2 outbreaks in China in Pudong, Shanghai and Jilin, Jilin. We estimated that the empirical effective reproduction numbers prior to the lockdown were 2.73 (95% CI: 2.42 – 3.05) for Pudong, and 2.81 (95%CI: 2.07-3.81) for Jilin (Supplementary Fig. 3), in agreement with our model estimates at baseline NPI intensity (Level 0). After implementing the lockdown, cases have been declining in both locations (Supplementary Fig. 3).

**Fig. 1.**
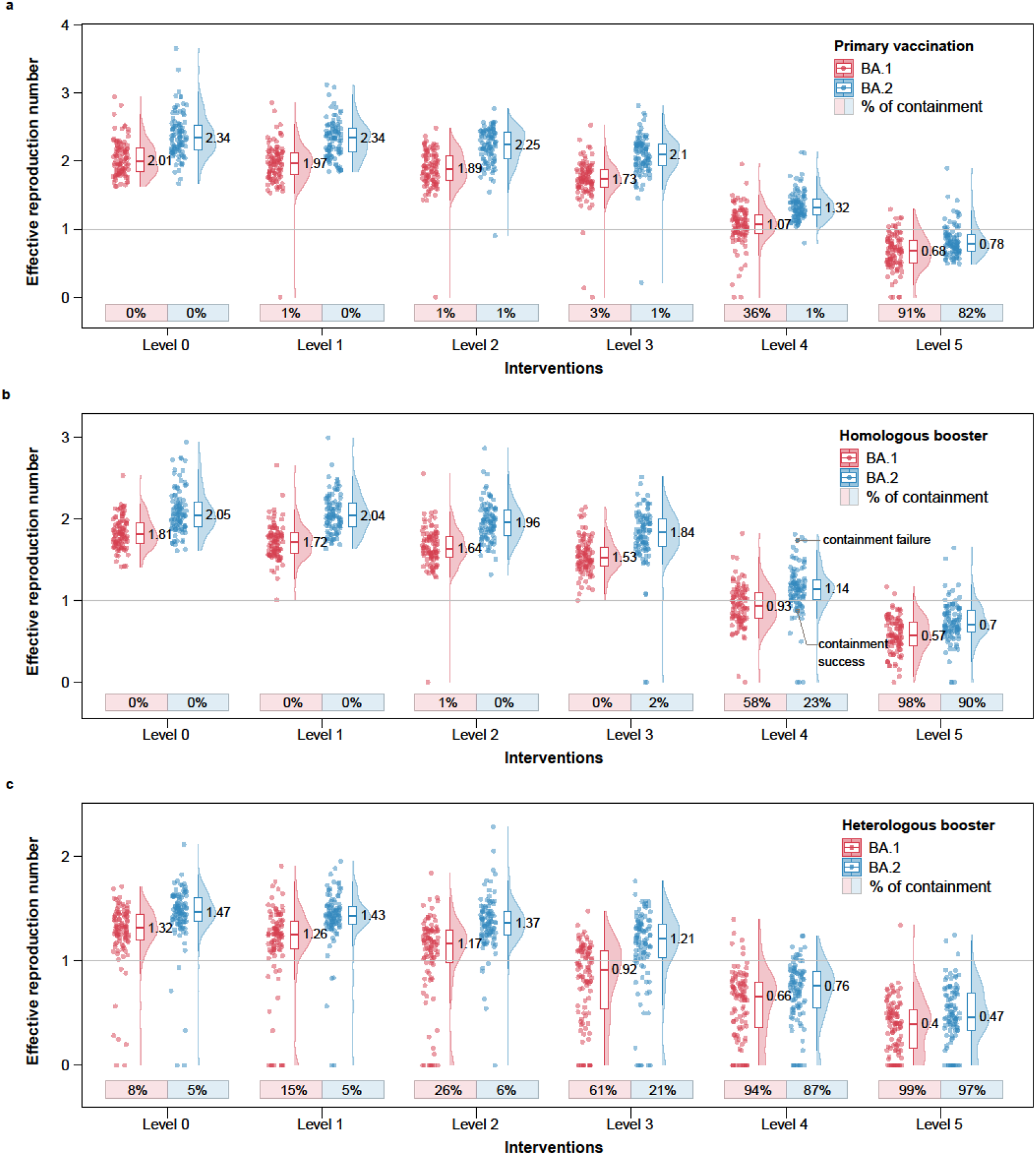
Likelihood of containing SARS-CoV-2 Omicron BA.1 and BA.2 outbreaks. **a**, Effective reproduction numbers and the probabilities of achieving containment under primary vaccination scenario and different level of NPI intensity, based on 100 simulations of the transmission chain. **b**, Same as **a** but for homologous boosting scenario. **c**, Same as **a** but for heterologous boosting scenario.

In the homologous boosting scenario (Fig. 1b), vaccination has only a moderate effect on preventing infection and reducing onward transmission, due to Omicron’s immune evasive properties against antibody neutralization activities ^15^ (Supplementary Table 1). Consequently, the average effective reproduction numbers under baseline NPI intensity are 1.81 for BA.1 and 2.05 for BA.2, both well above the epidemic threshold (Fig. 1b, Level 0). However, these booster strategies result in significant reductions in rates of severe illness and death, as demonstrated in earlier work ^8^. The high effective reproduction number under the baseline NPI intensity demands more stringent NPI measures for outbreak containment. Our modeling results suggest that even at Level 4 NPI intensity (with enhanced mass testing, Table 1), the average effective reproduction number hovers around the epidemic threshold of 1 (i.e., *R*_*eff*_=0.93 for BA.1 and *R*_*eff*_=1.14 for BA.2). As a result, the outcome of a containment effort becomes highly stochastic with approximately even chances of containment success vs. failure. In the case of containment failure, temporary population level mobility restrictions need to be swiftly implemented (Fig. 1b, Level 5) to bring the outbreak to an end.

As heterologous boosters with stronger immune response (relative to homologous boosters) were approved by the Chinese authorities in February 2022 ^16^, we explore the conditions for outbreak control with full coverage using a heterologous booster (Fig. 1c). Under the baseline NPI intensity, and despite full booster coverage of the entire population, vaccine breakthroughs will occur due to Omicron’s immune evasion properties (Supplementary Table 1). The outbreak is unlikely to be contained, with *R*_*eff*_=1.32 for BA.1 and *R*_*eff*_=1.47 for BA.2 (Fig. 1c, Level 0). However, heterologous boosters increase the population immunity against SARS-CoV-2 transmission, relative to homologous boosters, making epidemic control more likely with less stringent interventions. Enhancing routine interventions (Fig. 2b) would incrementally reduce transmissibility, with the probability of containment already over 50% at Level 3 intervention intensity for BA.1. For BA.2, probability of containment is lower (21%), suggesting that more containment efforts are needed to control this more transmissible lineage. Layering enhanced mass testing could achieve approximately 90% containment success for both BA.1 and BA.2, making population-level lockdowns unnecessary for most scenarios (Fig. 1c, Level 4 to 5).

**Fig. 2.**
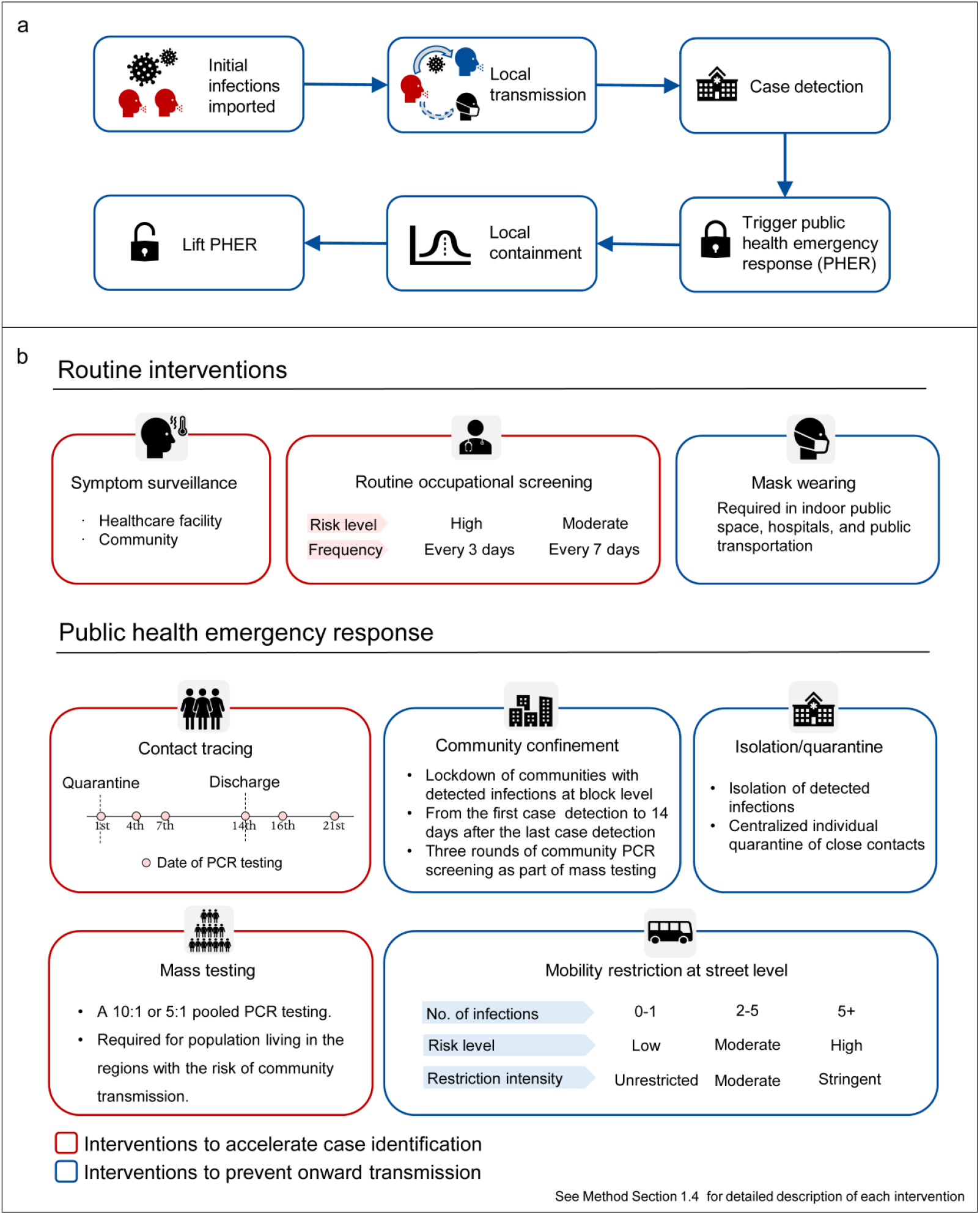
Non-pharmaceutical interventions to contain COVID-19 outbreak in Beijing, China. **a**, Public health response after the virus introduction. **b**, Routine interventions and public health emergency response (PHER) to accelerate case identification and prevent onward transmission.

In Fig. 3, we focus on two individual simulations of Omicron BA.2 outbreaks (annotated on Fig. 1b) under homologous booster scenario and Level 4 NPI intensity, representing containment failure (Fig. 3 a-c) and containment success (Fig. 3 d-f) respectively. The containment failure situation is characterized by growing incidence over time, spatially dispersed distribution of infections, and effective reproduction number above the epidemic threshold. On the contrary, containment success is characterized by a clear inflection point of the incidence curve, a localized outbreak, and an effective reproduction number swiftly dropping below the epidemic threshold. This illustrates how epidemiologic indicators of the trajectory of incidence and *R*_*eff*_ need to be closely monitored to guide decisions on the necessity of imposing stricter interventions. Temporary implementation of enhanced mobility restrictions and/or population-level lockdowns can allow for other, more targeted NPI measures to catch up and synergistically contain the spread of highly contagious variants.

**Fig. 3.**
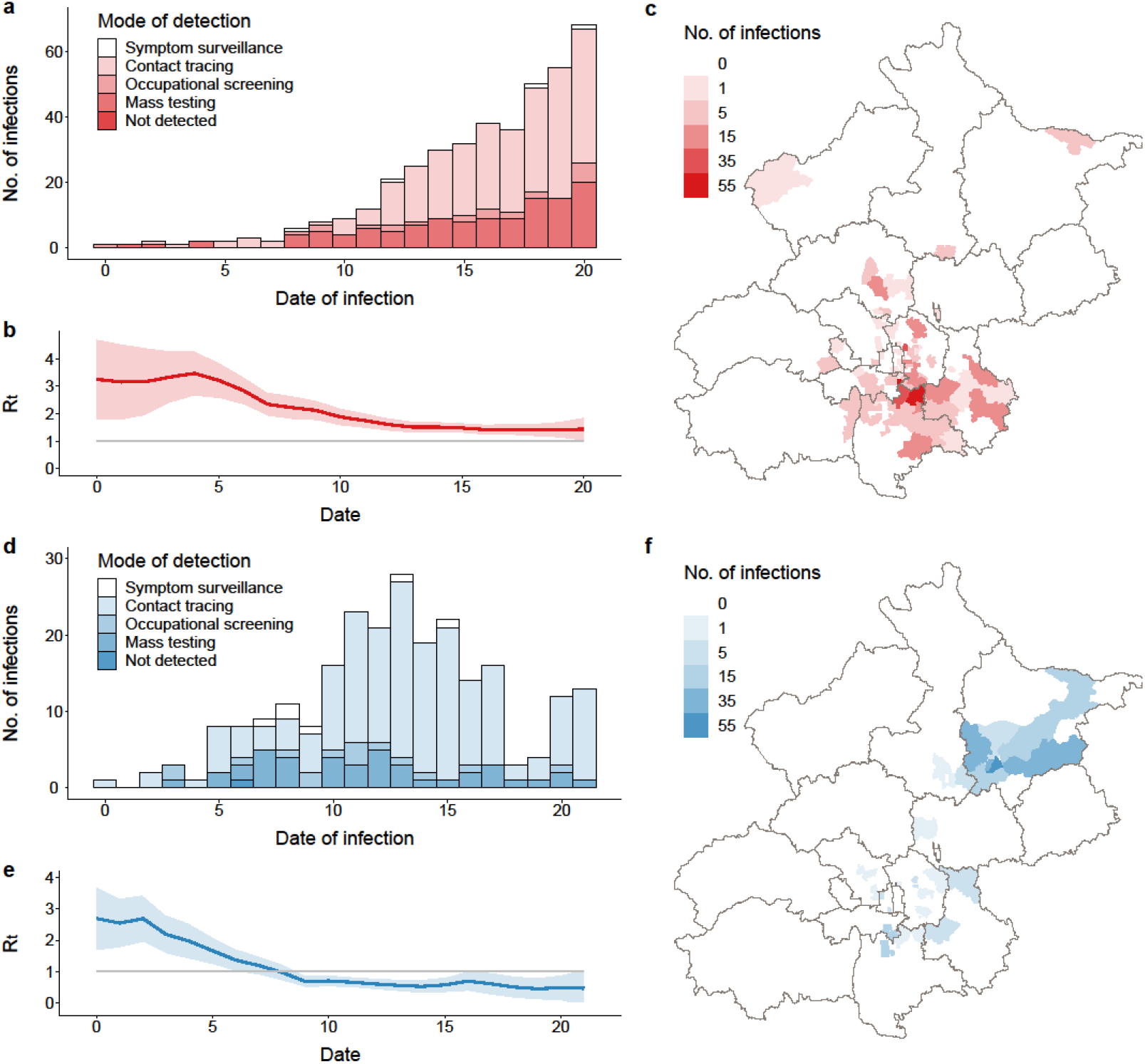
Individual simulations representing situations of containment failure and containment success of an Omicron BA.2 outbreak, under homologous booster scenario, with Level 4 NPI intensity. **a**, Daily number of new infections in an uncontained outbreak, stratified by mode of detection. **b**, the 5-day moving average effective reproduction number in an uncontained outbreak. The shaded area represents the 95% confidence interval. **c**, Spatial distribution of infected individuals in in an uncontained outbreak. **d-f**, Same as **a-c** but for a contained outbreak.

Finally, to explore the feasible pathways for mainland China to transit from SARS-CoV-2 local containment to mitigation, we built a statistical model to estimate the overall disease burden (measured in the total number of deaths) under various hypothetical mitigation scenarios across different levels of vaccination coverage, antiviral uptake, and SARS-CoV-2 variant types. The model, detailed in the Methods Section 3, relied on the empirical observations of the disease burden of Hong Kong’s Omicron BA.2 wave across different age-groups and vaccination status. For each mitigation scenario, we estimated 1) a “short-term” burden of one epidemic wave, assuming 60% of the population infected, similar to the epidemic size of the Hong Kong BA.2 wave, and 2) a “long-term” burden for the primary infections, assuming 95% of the population have been infected at least once, while neglecting burdens of subsequent repeat infections. As of March 17, 2022, although mainland China have reached an overall primary vaccination coverage of 87.9% and booster coverage of 45.7%, vaccine coverage in the older population significantly lagged behind that of the general population, with 50.7% of people over 80 years old covered by the primary schedule and 19.7% covered by the booster (Supplementary Fig. 2). For the Omicron’s BA. 2 sublineage (baseline variant), we estimated a total of 0.75 million deaths for the short-term burden and 1.18 million deaths for the long-term burden of primary infections, if China were to reopen under the vaccine coverage as of March 17, 2022, and with no uptake of antiviral treatment (the reference scenario, Fig. 4). Importantly, even though unvaccinated people only account for 9.7% of the total population in mainland China, they are responsible to 72% - 77% of the disease burden. However, through catching up on immunization in the older population and reaching high vaccine coverage across all age groups (same as that of New Zealand, Supplementary Fig. 2; high vaccine coverage scenario for Omicron BA.2, Fig. 4), both the short-term and long-term disease burden could be drastically reduced by 74.61%. Topping high vaccination coverage with increasing antiviral uptake could further reduce the disease burden: assuming 70% of the patients who are at risk of severe outcomes could be treated timely with antiviral drugs with 90% effectiveness of preventing deaths ^17^, we estimated a 90.61% reduction in disease burden comparing to that of the baseline scenarios, with 0.07 million deaths for the “short term” burden, comparable to that of the annual disease burden of seasonal influenza in mainland China ^18^. For sensitivity analysis, we assume that the age and vaccination status distributions among infections are the same as that of the entire population (while the main analysis assume that the age and vaccination status distributions among infections are the same as that of the observed infections in Hong Kong, detailed in Methods Section 3). The sensitivity analysis provides upper bound of projections, with 1.11 million and 1.75 million deaths for the short-term and long-term burden of primary infections in the reference scenario. Topping high vaccination coverage with 70% antiviral uptake could substantially reduce the disease burden by 93.20% (Supplementary Fig. 4). These estimates were also in agreement with the projections of Cai et. al., based on the projections of dynamical models ^14^. Given that SASR-CoV-2 has been continuously undergoing rapid adaptive evolution ^19^, we further consider a hypothetical SARS-CoV-2 variant with 50% increase in disease severity and another one with increased immune evasive properties comparing to Omicron BA.2 (detailed description in Supplementary Table 2). Both variants would lead to substantially higher burden when comparing to BA.2, but with high vaccination coverage (same as that of New Zealand, Supplementary Fig. 2) and antiviral drugs uptake (70%), the burden can still be effectively reduced to very tolerable levels: only 14.09% and 16.39% of the disease burden of the reference scenario (an Omicron BA.2 wave, Fig. 4). Overall, reaching high vaccination coverage across all age groups and stockpiling adequate among of effective antivirals are two of the most necessary conditions for China to transit from SASR-CoV-2 local containment into mitigation while avoiding high mortality burden.

**Fig. 4.**
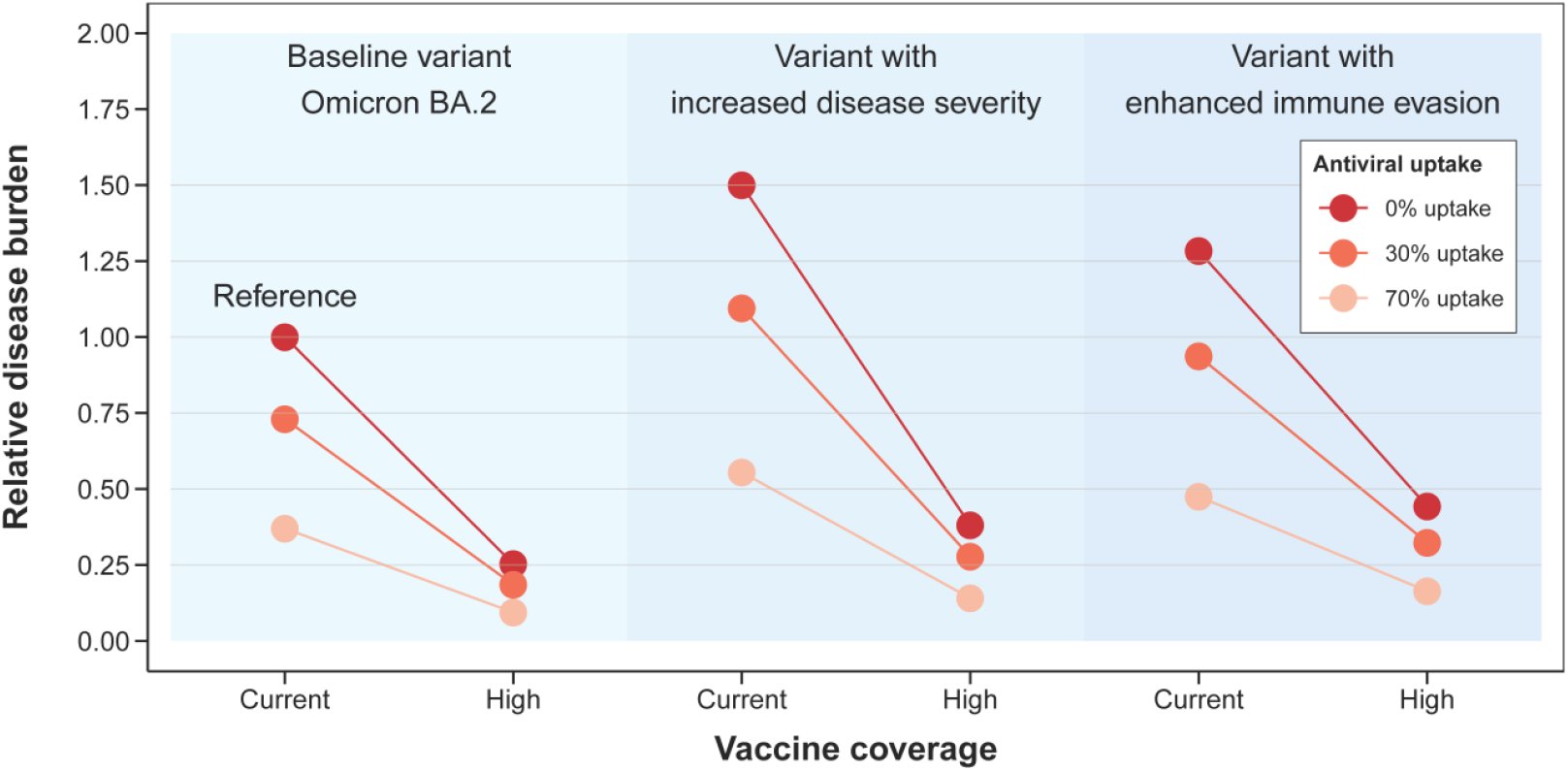
Relative disease burden of SARS-CoV-2 under different counterfactual mitigation scenarios in mainland China. Assuming the hypothetical situation that China have transitioned from SARS-CoV-2 containment to mitigation, we estimated the SARS-CoV-2 disease burden under different vaccine coverage and antiviral uptake levels. We consider SASR-CoV-2 BA.2 as the baseline variant, but also consider two hypothetical variants with increased disease severity/enhanced immune evasion properties relative to BA.2. The detail definition for each scenario is listed in Supplementary Table 2. The overall disease burden is measured in total number of deaths in mainland China. We consider the reference scenario as reopening the country with Omicron BA.2 introduced to mainland China under the vaccination coverage on March 17, 2022 and with no antiviral uptake. We provide relative disease burden of each scenario comparing to that of the reference scenario based on the lower bound projections estimated from a statistical model (detailed in Methods Section 3) based on the outcome the Omicron wave in Hong Kong SAR, China.

## Discussion

In this study, we utilized a detailed model of SARS-CoV-2 transmission and intervention, informed by age-specific contact patterns and high-resolution mobility data, to evaluate the feasibility of containment of Omicron’s BA.1 and BA.2 sublineages in the urban setting of Beijing. We explored an array of vaccine coverages and booster strategies to reflect existing and future population immunization levels. The demographic and spatial resolution of the model allowed us to characterize each non-pharmaceutical intervention explicitly and to dissect the contribution of each intervention individually. The stochastic nature of the model, together with consideration of individual transmission heterogeneity, enabled a probabilistic evaluation of the likelihood of outbreak containment. An interesting observation stemming from our model is that the overall effective reproduction number fluctuates drastically across simulations within the same scenario, reflecting the intrinsic stochasticity of SARS-CoV-2 transmission. Such observations have important real-world implications: under a given NPI intensity, a few anecdotal successes of containment for a specific variant does not guarantee that such NPI strategy will always be successful in the future. The containment of an outbreak could partially be due to stochastic fadeouts. A contingency plan to heighten NPI intensity needs to be ready to deploy swiftly.

Among all the analyzed NPI strategies, enhanced mass testing—increasing testing frequencies and expanding the size of the tested population—had the biggest effect on reducing transmission, besides population level mobility restrictions. The effectiveness of population-wide testing in reducing SARS-CoV-2 community transmission has also been demonstrated elsewhere ^20^. If contact tracing efforts are unable to identify epidemiological links between the detected infections, then enhanced mass testing of the population in high-risk regions needs to be swiftly implemented to uncover unobserved infections and interrupt transmission chains while the outbreak is still geographically limited. Logistically, enhanced mass testing would require a ramp-up of capacity over a short period of time. Building technical and logistic capacity for PCR testing before outbreaks materialize is of the highest priority to prevent Omicron and any future variant epidemics. However, this does not imply that other enhanced NPI measures—such as enhanced symptom surveillance, enhanced mask wearing, and enhanced occupational screening—are unimportant. All NPIs contribute incrementally to reducing SARS-CoV-2 transmission and could help elevate intervention pressures during the initial ramp up of mass testing. The additional transmission reduction effects of other NPI measures also contribute to faster outbreak suppression, decreased death and hospital burden, and faster return to normality. In addition, due to the stochastic nature of SARS-CoV-2 transmission as well as Omicron’s high transmissibility, targeted interventions cannot always contain outbreaks even with high vaccination coverage. The epidemic trajectory needs to be closely monitored throughout the entire course of an outbreak and stricter interventions such as temporary city-wide lockdown should be immediately implemented once rapid growing incidence and/or geographical dispersion are observed. Missing the optimal time of intervention will result in high caseloads that could overwhelm testing and contact tracing capacity, and in turn lead to reduced effectiveness of targeted NPIs and faster epidemic growth, akin to the situation of the on-going (as of April 27 2022) outbreaks in Shanghai and Jilin which have forced these cities into prolonged lockdown ^21^, illustrating that there is a short window of opportunity for interventions.

When facing the immune evasive Omicron variants, both the homologous and heterologous booster vaccination programs have only a marginal effect in reducing transmission and are thus inadequate to achieve epidemic control even when accompanied by the baseline non-pharmaceutical interventions. However, studies have shown that while all currently available vaccines are only partially effective in preventing infection, the vaccine effectiveness against severe outcomes remains high following the administration of a booster shot ^2,8^. Boosting population immunity and filling vaccine coverage gaps, especially for frail subgroups at high risk of COVID-19, and stockpiling effective antiviral drugs are top priorities; in the interim, local containment policies should be sustained. Moreover, given waning of immunity, sustained viral circulation in human and animal populations, and the repeated emergence of new variants, long-term plans for COVID-19 control in the post-pandemic phase would likely need to consider moving away from elimination and favoring transmission mitigation and disease burden reduction. However, mainland China’s current vaccination gap in older age groups, akin to the situation in Hong Kong (Supplementary Fig. 2) ^10^, would not permit an immediate transition to mitigation without causing significant hospitalization and death burden ^14^. Sustaining local containment is important until a high enough vaccination coverage can be reached and a decoupling between infection and severe disease can be ensured. Recent experiences with high vaccine coverages and mild Omicron outbreaks in New Zealand and Singapore ^9^ illustrate how a safe post-pandemic exit strategy could operate in mainland China.

Our model has several limitations. First, vaccine effectiveness to prevent Omicron infection is calculated based on neutralizing antibody titers and the current established correlate of protection ^22^. However, the correlate of protection analysis was based on symptomatic disease with the ancestral strain, thus it may not apply to other variant or asymptomatic infections. We also didn’t consider waning of immunity, although our model assumes low effectiveness against Omicron infection to start with. The temporal span of our simulations is limited as well. Future field studies of the effectiveness of inactivated vaccines against Omicron for both primary and booster doses are needed. Second, as of April 2022, the currently available evidence for BA.2’s fitness advantage over BA.1 is limited. In this study, we assume BA.2 has the same immune evasion properties, but heightened transmissibility based on preliminary data from the UK ^23^. Further evidence could help refine scenarios and provide more realistic assessments. Third, our analysis is based on the properties of the Omicron BA.1 and BA.2 sublineages. However, new variants/sublineages with even better fitness advantages over currently circulating ones could emerge in the future. The feasibility of containment against future variants needs to be critically reassessed to inform policy decisions. Finally, we considered 100% vaccine coverage of primary courses and boosters, which is a high bar, and hence our estimates of containment should be deemed optimistic. We note that China has been successful in reaching very high vaccine coverage for other immunization programs ^24^.

Overall, our study shows that local containment policy is not viable with sole reliance on vaccination, but it can be sustained by implementation of increased and highly reactive NPI interventions. Early outbreak detection, monitoring of epidemic indicators, and proactive control (particularly mass testing) are key to limit outbreak diffusion before it is too late. A rapid increase of coverage of primary vaccine courses and boosters among high-risk population as well as stockpiling highly effective antiviral drugs are of utmost importance before China can exit out of the local containment phase and move into long-term mitigation phase while SARS-CoV-2 transitioning into endemicity.

## Methods

### 1 Evaluating the feasibility of containment strategy against Omicron variants, based on a spatially structured individual-based SARS-CoV-2 transmission model

#### 1.1 Overview

Here we use a spatially structured individual-based SARS-CoV-2 transmission model to evaluate the feasibility of containment strategy against Omicron BA.1 and BA.2 in a densely populated urban setting in China. In particular, the model built upon prior work that was applied to reconstruct the containment of the Xinfadi outbreak in Beijing caused by the ancestral SARS-CoV-2 lineage ^13^. The model was further expanded to incorporate Omicron-specific epidemiology, in terms of transmissibility, generation interval, and immune evasion properties. The model considers various levels of non-pharmaceutical intervention strategies to reflect enhanced outbreak response protocols that have been adopted in China over time. Finally, the model further incorporates up-to-date vaccination coverage to reflect the current (as of March 2022) and future level of population immunity in China.

#### 1.2 Individual reproduction number and SARS-CoV-2 transmission as branching process

To simulate the spread of SARS-CoV-2 in the absence of NPIs, we start by seeding the population of interest with three infected individuals. Each individual *i* infected at time *t*_*i*_ can either be classified as symptomatic or asymptomatic based on the age-specific asymptomatic rate ^25^ of SARS-CoV-2 *Φ*_*asymp*_. For symptomatic infections, we assign the time delay from infection to symptom onset *τ*_*incu*_ by drawing from the incubation period distribution *P*_*incu*_(*τ*) ^26^. To simulate the transmission of SARS-CoV-2 at the individual level, in the absence of NPIs, we assign individual *i*’s reproduction number *R*_*i*_ (number of secondary infections caused by *i*), by drawing from a negative binomial distribution *NB*(*R*_0_, *k*), where the mean of the negative binomial distribution *R*_0_ is the basic reproduction number (population average of *R*_*i*_), and *k* is the dispersion parameter of negative binomial distribution, capturing the heterogeneity of SARS-CoV-2 transmission (we provide *R*_0_ and *k* values specific for Omicron later in Methods Section 1.5). Therefore, individual *i* would cause a total of *R*_*i*_ secondary infections. We assume the shape of each infected individual’s infectiousness profile follows the distribution of the generation interval *GI*(*τ*). Thus, the timing of transmission *τ*_*ij*_ from individual *i* to individual *j* ∈ {*R*_*i*_} is given by *τ*_*ij*_ = *t*_*j*_ − *t*_*i*_, where *t*_*i*_ and *t*_*j*_ are the *i* and *j*’s timing of infection, and *τ*_*ij*_ is drawn from the generation interval distribution *GI*(*τ*). We recursively simulate the onward spreading of secondary infections through multiple generations until the 30^th^ day after virus introduction or the daily number of infections reaches 10,000.

#### 1.3 Population structure reflecting age-specific contact patterns, occupation, transmission setting, and spatially resolved mobility patterns

When transmission between primary infection *i* and their contact *j* occurs, we first generate the setting of the transmission event (home, workplace, or community) permissive by the occupation of the primary infector *i*. We then assign the age of secondary infection *j* based on the age-specific contact matrices and the transmission setting. We further probabilistically determine *j*’s occupation in accordance with their age based on the transmission setting and primary infector *i*’s occupation. Conditional on the transmission setting, the geographical locations of the residences of secondary infections were assigned based on the street/town level mobility network from aggregated mobile phone data provided by China Unicom, one of the leading mobile phone service providers in China (see the study by Wang et. al. ^13^for more details).

#### 1.4 Non-pharmaceutical interventions (NPIs)

The details of the public health measures in response to local outbreaks in China have been previously described ^27,28^. As shown in Fig. 2, we categorize these measures into seven types: (i) symptom-based surveillance in healthcare facilities and communities; (ii) mask-wearing order in public places; (iii) routine screening for workers with risk of occupational exposure; (iv) systematic tracing, quarantine, and testing of close contacts; (v) lockdown of residential communities with detected infections; (vi) mass testing and (vii) mobility restrictions based on regional risk levels. All intervention strategies are summarized in Fig. 2, of which symptom surveillance, mask wearing, and occupational screening are routine interventions implemented regularly, while contact tracing, community confinement, mass testing, and mobility restrictions are public health emergency response only conducted when new infections are detected. We modeled the impact of NPIs by emulating their effect on accelerating case detection and preventing new infections. Below we give a brief description of each NPI:

- **(i) Symptom surveillance:** Individuals who present symptoms consistent with SARS-CoV-2’s clinical presentation during healthcare consultations at hospitals and local clinics would be considered as SARS-CoV-2 suspected cases and be provisionally isolated in designated facilities. At least 3 subsequent PCR tests for SARS-CoV-2 diagnostics would be conducted at the 1^st^, 3^rd^, and 7^th^ day of isolation. If the suspected case is diagnosed by molecular tests, they would remain in hospital and be treated until the individual is fully recovered and no longer infectiousness. To heighten case detection through symptom surveillance, routine temperature checking is implemented in public spaces, such as workplaces, markets, shopping malls, subway stations, railway stations, and airports. Any individual with suspected symptoms of SARS-CoV-2 will be asked to seek medical attention.
- **(ii) Mask wearing:** Mask wearing is required in public spaces, including hospitals, public transportation, markets, shopping malls, and entertainment venues.
- **(iii) Occupational screening:** Routine PCR screening is conducted among individuals with occupations at risk of SARS-CoV-2 infection and/or in frequent contact with the general population, including: staff at customs and border control, international ports, and quarantine locations; healthcare workers; individuals working in confined environments (e.g., institutions providing long term care and prisons); staff providing public services (e.g., public transportation, delivery, museums, and libraries); and workers in businesses and wet markets. The screening frequency is determined based on the occupational exposure risk and the screening population can be adjusted according to the actual situation.
- **(iv) Contact tracing:** A close contact is defined as person who interacts with a confirmed or suspected COVID-19 case for the period from 4 days before to 14 days after the illness onset, or with an asymptomatic carrier for the period from 4 days before to 14 days after collection of the first positive sample. Close contacts are further grouped into household contacts, work contacts, and community contacts based on their transmission setting. Epidemiological investigations to identify close contacts, including manual investigations (e.g., phone contact, interview) and electronic tracing (e.g., mobile apps, online databases), should be completed within 24 hours. Centralized quarantine for at least 14 days is required for all close contacts with periodic PCR testing at the 1^st^, 4^th^, 7^th,^ and 14^th^ day of quarantine and the 2^nd^ and 7^th^ day after discharge.
- **(v) Residential community confinement:** The residential communities with detected infections are on lockdown at the building block level until 14 days after the last case identification, with stay-at-home orders for all residents other than essential workers. Supplies of living necessities are provided by community workers. Three rounds of PCR screening are conducted as part of mass testing for all community residents at the 2^nd^, 8^th^ and 13^th^ day of lockdown, respectively.
- **(vi) Mass testing:** Emergency mass testing in a targeted area is immediately activated when a new infection is reported. The geographical scope is usually determined according to the administrative division. Multiple rounds of testing are often conducted, and each round should be completed within 3 days. A 10:1 or 5:1 pooled sample approach is used to expand the PCR capacity and increase cost effectiveness.
- **(vii) Mobility restrictions:** Unrestricted movement is allowed in low-risk areas, i.e., streets/towns with zero to one detected infection. The street/town is upgraded to moderate risk once it has reported more than one infection, with entertainment venues being closed and mass gatherings being prohibited. Residents in moderate risk areas are required to avoid unnecessary travel. A street/town with more than five infections will be upgraded to high risk with more stringent population mobility restrictions being implemented, where all public transportation within and in and out of the area will be suspended. Temporal lockdown is sometimes adopted in high risk areas, with all residents except for essential workers staying at home during lockdown. The street/town will be downgraded to low risk if no new infections are reported for 14 consecutive days, with mobility restrictions gradually lifted.

The detailed implementations of each NPI into the transmission model has been previously described in the study by Wang et. al. ^13^. The strengths of each NPI in different scenarios considered are presented in Table 1.

#### 1.5 Omicron BA.1 and BA.2 sublineages

Current (April 2022) evidence suggests the Omicron variant has significant fitness advantage over Delta and has replaced Delta to become the dominate variant globally. A household transmission study from Denmark has shown that the Omicron BA.1 variant is 17% more transmissible than the Delta variant ^29^ for unvaccinated individuals, while the BA.2 variant is 27% more transmissible than BA.1 ^23^. Assuming that Delta’s basic reproduction number is 6.4 ^30^, then the basic reproduction numbers for BA.1 and BA.2 are 7.5 and 9.5 respectively. The generation interval, incubation period, overdispersion, and symptomatic proportion are assumed the same for BA.1 and BA.2. For unmitigated transmission, we assume Omicron variant transmission’s offspring distribution follows a negative binomial distribution with mean *R*_0_ and the dispersion parameter *k*00.43 ^31^. We assume the mean intrinsic generation interval of Omicron to be the same as Delta (4.7 days) ^32^. We assume the incubation period follows gamma distribution with a mean of 5.8 days and standard deviation of 3.0 days ^26^. We hypothesize that the proportion of symptomatic infections *Φ*_*symp*_ increases with age, with 18.1%, 22.4%, 30.5%, 35.5%, and 64.6% of infections (without vaccination) developing symptoms in groups aged 0-19, 20-39, 40-59, 60-79, and 80 or more years respectively ^25^.

#### 1.6 Vaccination coverage and population immunity

As of March 17, 2022, a total of 3.21 billion doses of SARS-CoV-2 vaccines have been administered in mainland China, including two inactive vaccines (CoronaVac and Covilo) and a viral vector vaccine (Convidecia) ^12^. As of March 17, 2022, the vaccine coverage of the primary series is 87.9%, on top of which 45.7% of the population have received a booster shot ^12^. The effectiveness of primary vaccination, homologous booster and heterologous booster in preventing infection (*VE*_*I*_), symptomatic disease (*VE*_*s*_) and onward transmission (*VE*_*T*_) caused by the Omicron variants was taken from the estimates in the study by Cai et. al. ^14^, either extracted from real-world studies or predicted based on neutralizing antibody titers (NATs), and summarized in Supplementary Table 1.

Since the containment strategy has been maintained in mainland China since the beginning of the pandemic, the proportion of the population who has been infected with SARS-CoV-2 remains extremely low nationally ^27,28,33^. In this study, we ignore the effect of infection-induced immunity and focus on vaccine-induced immunity.

#### 1.7 Scenarios of SARS-CoV-2 containment under different variants, levels of population immunity, and NPI strengths

To anticipate the feasibility of maintaining the SARS-CoV-2 local containment in mainland China in 2022, we evaluate an exhaustive combinatory of hypothetical scenarios along the dimensions of different Omicron sublineages, level of population immunity, and strength of NPIs.

##### Variant type

We consider both the Omicron BA.1 and BA.2 sublineages, as they account for the vast majority of the currently circulating variants. According to prior studies, Omicron BA.1 is 17% more transmissible than the previously circulating Delta variant, with the corresponding basic reproduction number *R*_0_ = 7.5 ^29^, while Omicron BA.2 has an increased transmissibility of 27% comparing to BA.1, with *R*_0_ = 9.5 ^23^.

##### Level of population immunity

We consider three immunization scenarios for the modelling analysis with the baseline immunization scenario assuming 100% coverage by primary series with 0% booster coverage and two enhanced immunization scenarios, one with 100% homologous booster coverage and the other with 100% heterologous booster coverage. As vaccine rollout continues in China, the real-world situation lies between the two enhanced immunization scenarios. The effectiveness of vaccination on preventing infection, symptomatic disease, and reducing onward transmission is described in section 2.5.

##### Intervention strategy

To assess the effectiveness of different containment strategies, we firstly set a baseline scenario with moderate intervention intensity. The NPI intensity is then gradually increased in the subsequent scenarios until we have enough confidence to achieve epidemic control. Under each scenario, we consider the following hypotheses: i) infections are isolated immediately at the time of laboratory confirmation; ii) isolation and quarantine are completely effective on preventing onward transmission; iii) the sensitivity of PCR testing varies with time, following the estimates of the prior study ^34^; iv) PCR testing should be completed (from collection of samples to reporting of results) within six hours ^28^; v) The protective effect of mask wearing against onward transmission and infection of SARS-CoV-2 is 9.5% and 18%, respectively ^35,36^; vi) all household contacts are immediately quarantined, while all work contacts and 70% of community contacts are quarantined with a mean time delay of 0.7 days. Details of each intervention scenario are described below. The intervention parameters are summarized in Table 1.

- **Level 0 (Baseline interventions):** In our baseline scenario, we hypothesize that 33.3% of the symptomatic infections seek healthcare attention with an average delay of 3.7 days after symptom onset. We assume a moderate mask-wearing order, with 10% of individuals wearing masks in the workplace and 30% in the community. For occupational screening, we assume that high-exposure risk population (i.e., relative risk (RR) 0 8 comparing to low-risk groups, 2.5% of the working-age population) are tested every 3 days; moderate-exposure risk population (i.e., RR02, 7.5% of the working-age population) are tested every 7 days. Emergency response, including contact tracing, community confinement, mass testing, and mobility restrictions, is triggered right after the identification of the first infection. Mass testing is conducted at the street/town level and must be completed within three days after the first infection being detected. Stringent or moderate mobility restrictions are implemented according to the risk level of each street/town based on the real-time assessment of local transmission risk. The hypothetical origin-destination mobility matrix depending on risk levels, is shown in Supplementary Table 3.
- **Level 1 (Level 0 + Enhanced symptom surveillance):** Comparing to the baseline intervention scenario, we enhance the intensity of symptom surveillance, with the proportion of detectible symptomatic infections increased from 33.3% to 66.7%, and the mean time delay from symptom onset to hospitalization shortened from 3.7 days to 2.7 days.
- **Level 2 (Level 1 + Enhanced mask wearing):** Comparing to Level 1 intervention scenario, a more stringent mask-wearing order is considered, assuming 50% and 80% of individuals wearing masks in the workplace and in the community, respectively.
- **Level 3 (Level 2 + Enhanced occupational screening):** Comparing to Level 2 intervention scenario, the population of occupational screening is then expanded, with 5% of working-age populations (i.e., RR= 7.5) included in high-risk group and tested every 3 days, and 20% (i.e., RR=1.875) included in moderate-risk group and tested every week.
- **Level 4 (Level 3 + Enhanced mass testing):** Comparing to Level 3 intervention scenario, we further enhance the intensity of mass testing, with the rounds of testing increased from 1 to 5, and the geographical range of testing expanded from the residential street/town of the detected infections to the whole district/county.
- **Level 5: (Level 4 + Enhanced mobility restrictions):** Comparing to Level 4 intervention scenario, mobility restrictions are further enhanced if the outbreak cannot be contained through previous efforts. In addition to the lockdown of high-risk regions, strict and moderate mobility restrictions are implemented in moderate-risk and low-risk regions, respectively. The hypothetical origin-destination mobility matrix depending on risk levels is shown in Supplementary Table 4.

We exhaustively explored all combinatory of two Omicron sublineages, three population immunity levels, and six NPI intensity levels, for a total of 36 scenarios.

#### 1.8 Outbreak Simulation

For each of the 36 scenarios described in the previous section, we seed outbreaks with three initial infections distributed according to population density ^37^. Their ages and occupations are sampled from the demographical structures ^38^. With variant-specific epidemiologic parameters and immunity induced infection prevention and transmission reduction, we first simulate the transmission chain in the absence of NPIs, based on a branching process described in Section 2.1. Even for the enhanced immunization scenario, all variants considered have effective reproduction numbers larger than 1, above the epidemic threshold, and are able to sustain transmission in the population. We simulate the transmission chain until either reaching the 30^th^ day after the virus introduction or when the daily number of new infections exceeds 10,000. We then simulate the effect of NPIs through pruning the unmitigated chains of transmission by removing branches that would otherwise be interrupted by the corresponding NPIs of the scenario of interest. Details of the implementations of the simulation can be found in the study by Wang et. al. ^13^. We run 100 simulations for each scenario to capture the stochasticity of the transmission process. For each simulation, the following summary statistics are calculated: i) the overall effective reproduction number (*R*_*eff*_), defined as the average of the individual reproduction number of each infection infected after the implementation of NPIs; ii) daily number of new infections by modes of detection; iii) The five-day moving average effective reproduction number (*Rt*) at day *t*, defined as the average individual reproduction number for a cohort of infections infected within the time window of day *t* till day *t* + 5; iv) the spatial distribution of SARS-CoV-2 infections at the street/town level. The branching process model is coded in Python 3.10. The statistical analyses and visualization are performed using R software, version 4.0.2.

### 2 Estimating effective reproduction number during the early phase of the Omicron BA.2 outbreaks in Pudong, Shanghai and Jilin, Jilin

In Supplementary Fig. 3, we plotted the daily incidence of the ongoing Omicron BA.2 outbreaks in Pudong District, Shanghai and Jilin, Jilin Province. The vertical dashed vertical lines indicate the timing of imposing lockdown in both locations. We estimated the epidemic growth rate before the lockdown (assuming exponential growth during this period) through fitting a linear regression to the incidence curve (in log scale) prior to the lockdown. The growth rate along with its uncertainties were estimated as the slope of the linear regression. Based on the estimated growth rates and the generation interval distribution (Methods Section 1.5), we estimated the effective reproduction number in Pudong and Jilin based on method proposed by Wallinga et. al. for empirical generation interval distributions ^39^:

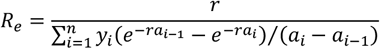

Where *R*_*e*_ denotes the effective reproduction number, *r* denotes the growth rate, *y*_*i*_ denotes the relative frequency of the histogram of the discretized generation interval at daily resolution and *a*_*i*_ denotes the category bounds in such histogram.

### 3 Projection of the SASR-CoV-2 disease burden in mainland China under different hypothetical scenarios, based on the observed disease burden of the Omicron BA.2 wave in Hong Kong SAR, China

As of May 11, 2022, the Omicron BA.2 wave in Hong Kong, China have subsided in terms of SARS-CoV-2 infections and deaths (Supplementary Fig. 1a) ^40,41^. The government of the Hong Kong Special Administrative Region also reported detailed morbidity and mortality data of the Omicron wave stratified by age and vaccination status ^42,43^. The Laboratory of Data Discovery for Health at the University of Hong Kong used mathematical model to project the epidemic trajectory and final epidemic size including all infections not limited to those have been captured by the surveillance system ^10^. Here we synthetize this information together and provide two bounding estimates of the infection fatality ratio for each of the specific age group and population with a given infection status. Specifically, let’s denote *c*_*αv*_ and *d*_*αv*_ as the number of reported SASR-CoV-2 infections and deaths of age bracket *α* and vaccination status *v*, where *α* could take the value of <3 years, 3-19 years, 20-39 years, 40-59 years, 60-69 years, 70-79 years, and 80 yeas and older and *v* could take the value of “Unvaccinated”, “CoronaVac 1 dose”, “CoronaVac 2 doses”, “CoronaVac 3 doses”, “Comirnaty 1 dose”, “Comirnaty 2 doses”, “Comirnaty 3 doses”, respectively ^43^. We could calculate the crude case fatality ratio *CFR*_*αv*_by age *α* and vaccination status *v* as:

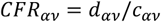

Supplementary Fig. 1c plotted the age-specific case fatality ratio by vaccination status for the Omicron BA.2 wave in Hong Kong. Unvaccinated individuals have the highest case fatality ratio across all age groups while individual who received 3rd doses of either of the CoronaVac or the Comirnaty vaccines have the lowest. However, it’s unlikely that Hong Kong’s surveillance system were able captured all infections among the Hong Kong population during the Omicron wave. Modelling analysis matching the epidemic trajectory of the Omicron wave have projected 4.4 million out of the 7.4 million Hong Kong population representing 60% infection attack rate. In the meanwhile, there are only 1.2 million reported infections as of May 11, 2022, suggesting roughly 1 in 4 Omicron BA.2 infections were reported. If we denote *i*_*αv*_ as the number of total infections of age bracket *α* and vaccination status *v*, then we can express the infection fatality ratio of the corresponding population:

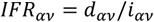

However, *i*_*αv*_ was not directly observed. To overcome this, we first assume that the total number of infections is the same as model projected 4.4 million, i.e.:

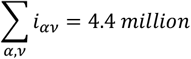

Under one extreme scenario, we assume that the distribution of *i*_*αv*_ across age *α* and vaccination status *v* is the same as the distribution of *c*_*αv*_ across age *α* and vaccination status *v*. i.e., *i*_*αv*_ ∝ *c*_*αv*_. Given that Σ_*α,v*_ *i*_*αv*_ = 4.4 million, we can calculate 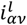 within age group *α* and vaccination status *v*, where *l* denotes this bounding scenario. The age and vaccination specific infection fatality ratio under this scenario *l* can by estimated as:

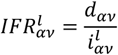

The Hong Kong Government also reported daily number of populations who had received 1^st^, 2^nd^ and 3^rd^ dose by vaccine type and age group *α* prior to the Omicron wave (February 15^th^, 2022) ^43^, based on which we could calculate the population size *p*_*αv*_ of age bracket *α* and vaccination status *v*. Under another extreme, we assume that the distribution of *i*_*αv*_ across age *α* and vaccination status *v* is the same as the distribution of *p*_*αv*_ across age *α* and vaccination status *v*. i.e., *i*_*αv*_ ∝ *p*_*αv*_. Given that Σ_*α,v*_ *i*_*αv*_ = 4.4 million, we can calculate 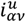 within age group *α* and vaccination status *v*, where *u* denotes this bounding scenario. The age and vaccination specific infection fatality ratio under this scenario *u* can be estimated as:

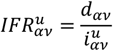

One issue is that we could not directly calculate *p*_*αv*_for the unvaccinated, thus we further assume that the fraction of population who got infected with 1 dose of the CoronaVac is the same as those who hadn’t received any vaccination. Thus, we could calculate *p*_*αv*_ for the unvaccinated as:

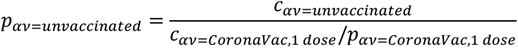

Then, we could estimate 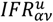 across all age groups and vaccination status, including the unvaccinated individuals. We assume that all vaccines (mostly inactivated) used in mainland China have the same vaccine effectiveness of CoronaVac. Give the infection rate, vaccination coverage, vaccination coverage, and the effects of antivirals listed in Supplementary Table 2, we could project the total number of SARS-CoV-2 caused deaths based on the estimated infection fatality ratio. Given that 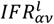 and 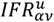 provides different estimates, and consequently different projections for the total number of deaths, we provide both 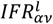 and 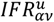 as the lower and upper bound of projections. The results of the lower bound projections are shown in Fig. 4. The results of the upper bound projections are shown in Supplementary Fig. 4, as a sensitivity analysis.

## Data Availability

All data produced in the present study are available upon reasonable request to the authors

## Data and code availability

All data and code to reproduce all figures in the main text and supplementary materials are publicly available upon acceptance of the manuscript.

## Acknowledgments

This study was supported by grants from the Key Program of the National Natural Science Foundation of China (82130093). The findings and conclusions in this report are those of the authors and do not necessarily represent the official position of the NIH.

## Author contributions

H. Y. designed and supervised the study. Y. Wang and K. S. carried out development of the model, designed the simulations, performed statistical analysis and drafted the manuscript. Z. F., L. Y., and Y. Wu. helped with constructing the figures. H. L. helped with inferring the effective reproduction number using the epidemic data. Q.W., M.A., C.V., and H.Y. made critical revision of the manuscript for important intellectual content. All authors were involved in the research and revising of the manuscript. All authors approved the final manuscript.

## Competing interests

H.Y. received research funding from Sanofi Pasteur, GlaxoSmithKline, Yichang HEC Changjiang Pharmaceutical Company, Shanghai Roche Pharmaceutical Company, and SINOVAC Biotech Ltd. Except for research funding from SINOVAC Biotech Ltd, which is related to the data analysis of clinical trials of immunogenicity and safety of CoronaVac, the others are not related to COVID-19. All the other authors have no competing interests.

## Supplementary information

**Supplementary Fig. 1.**
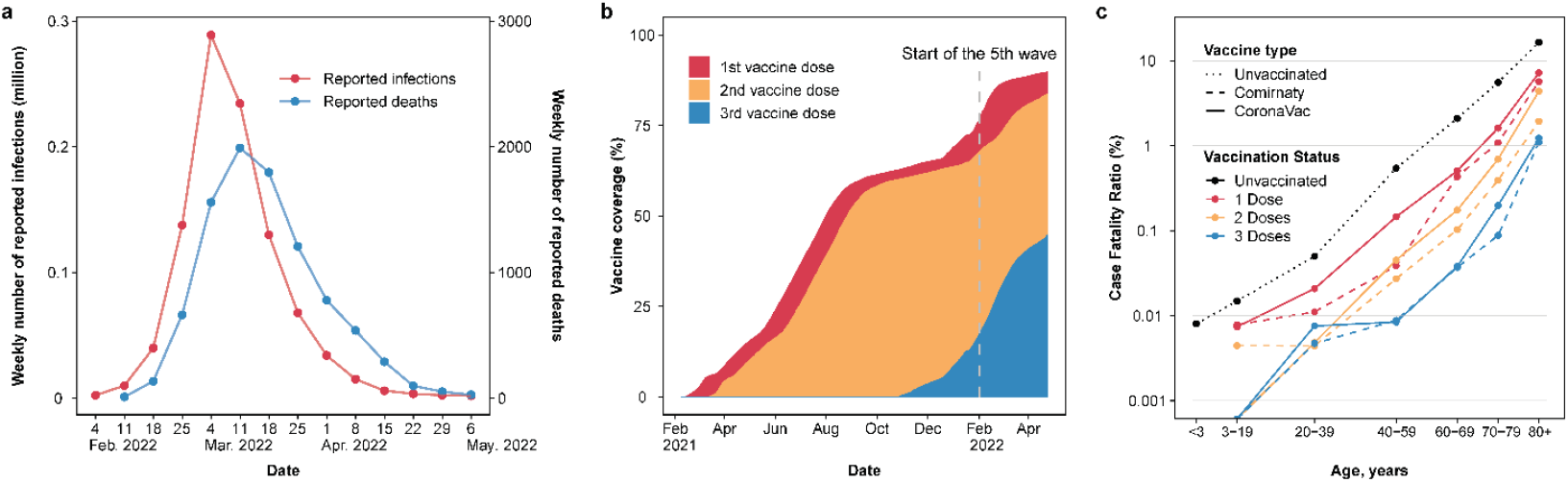
Epidemic trajectory, vaccine uptake, and clinical severity of Hong Kong SAR’s Omicron wave. **a**, Weekly incidence of reported Omicron infections and deaths from February 1 to May 9, 2022 ^1,2^. **b**, Vaccine coverage in Hong Kong until May 8, 2022 stratified by vaccine dose ^3^. **c**, Hong Kong’s crude case fatality ratio by age and vaccine status ^4^.

**Supplementary Fig. 2.**
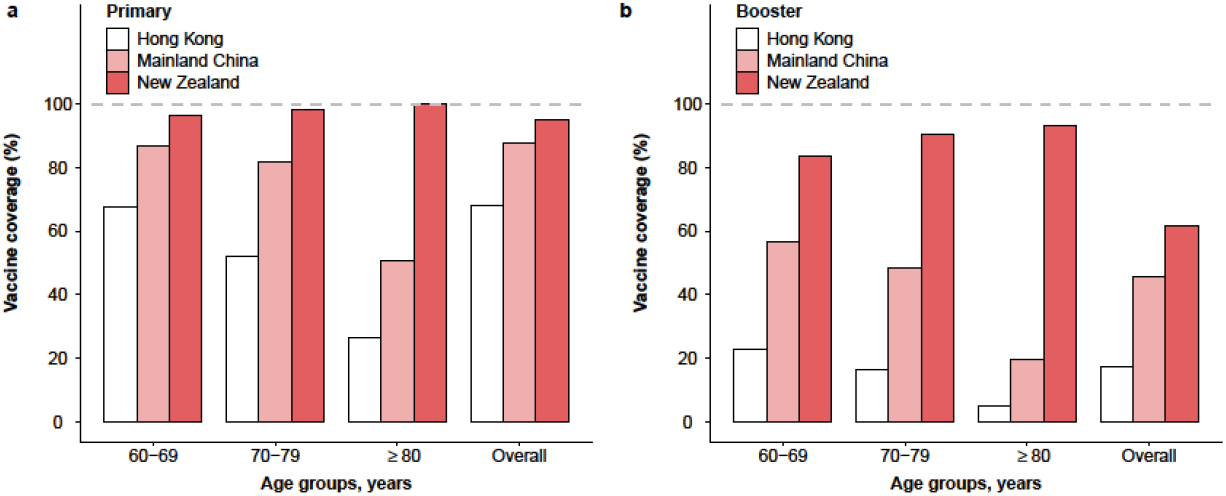
Vaccinations uptake by age in Hong Kong SAR, China (as of February 15, 2022), Mainland China (as of March 17, 2022) and New Zealand (as of April 5, 2022) ^3,5,6^. **a**, Primary vaccination. **b**, Booster vaccination.

**Supplementary Fig. 3.**
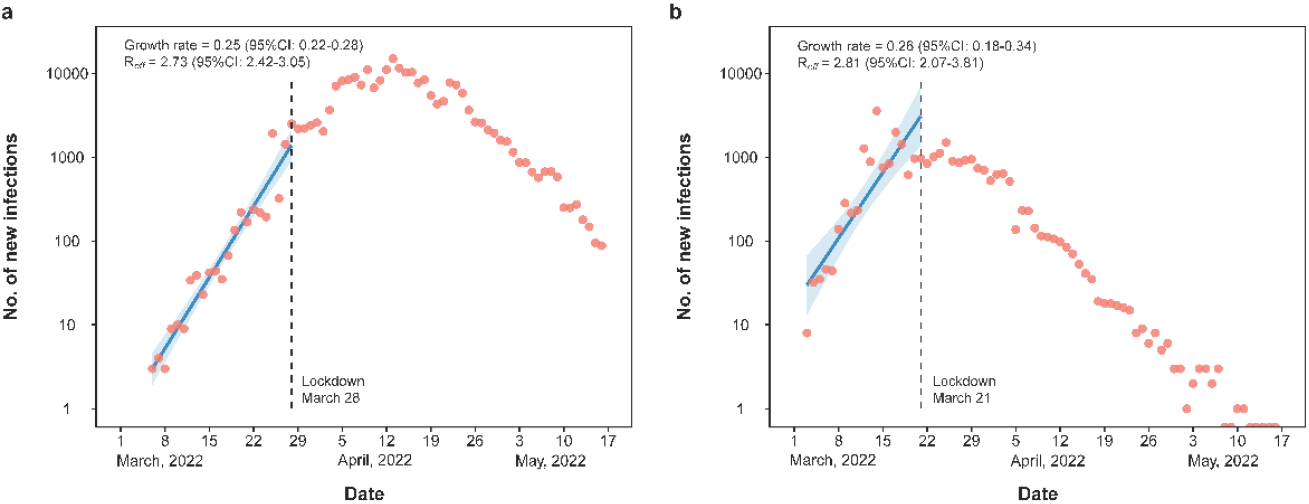
The observed epidemic growth rate and effective reproduction numbers with baseline interventions in the early stage of the two recent outbreaks in mainland China. **a**, the outbreak in Pudong new district, Shanghai. Red dots are daily new infections and the blue line best fitted growth rate. The blue shade represents 95%CI. **b**, Same as **a** but for the outbreak in Jilin city, Jilin province.

**Supplementary Fig. 4:**
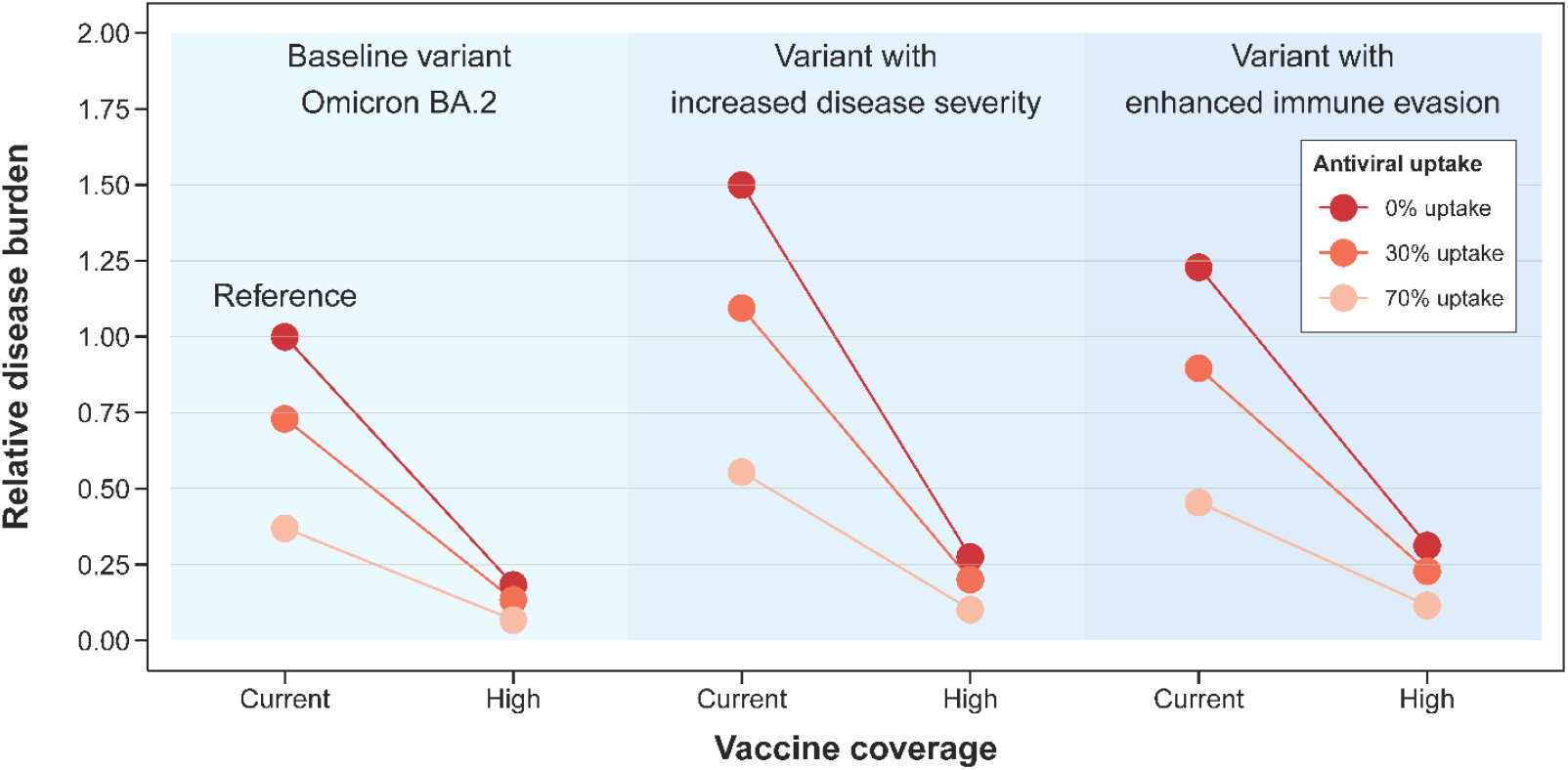
Relative disease burden of SARS-CoV-2 under different counterfactual mitigation scenarios in mainland China. As a sensitivity analysis, we provide relative disease burden of each scenario based on the upper bound projections. The results based on the lower bound projections are shown in Fig. 4.

**Supplementary Table 1.**
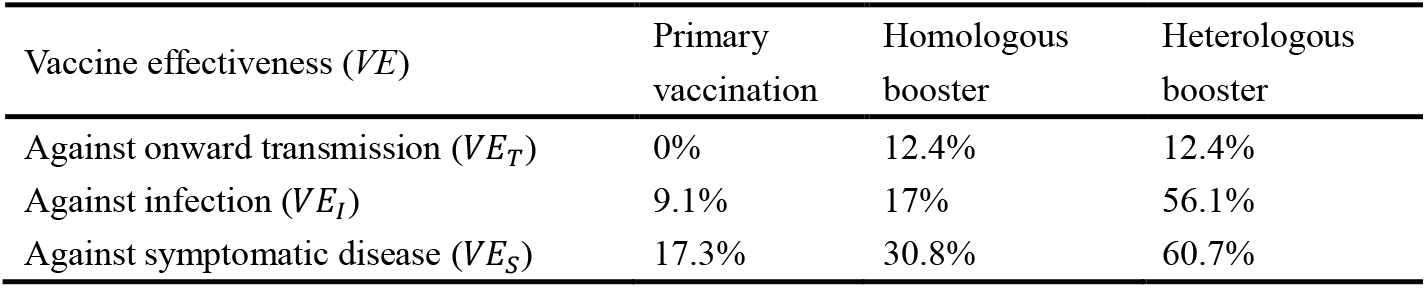
Effectiveness of COVID-19 vaccines.

**Supplementary Table 2.** List of considered hypothetical mitigation scenarios.

| Mitigation scenario |  | Definition |
| --- | --- | --- |
| Variant type | Baseline variant (Omicron BA.2) | The Omicron BA.2 variant with estimated age and vaccine status specific infection fatality ratios (IFR) estimated based on the Hong Kong BA.2 wave (Material and Methods Section 4). |
|  | Variant with increased disease severity | A hypothetical more severe variant with age and vaccine status specific IFRs 50% higher than those of the Omicron BA.2. |
|  | Variant with enhanced immune evasion | A hypothetical immune evasive variant with age specific IFRs the same as those of Omicron BA.2 for the unvaccinated individuals but 100% higher among vaccinated individuals. |
| Vaccine coverage | Current | The mainland China's age-specific vaccination coverage as of March 17<sup>th</sup>, 2022 (Supplementary Fig. 2) <sup>5</sup>. |
|  | High | The New Zealand's age-specific vaccination coverage as of April 5, 2022 (Supplementary Fig. 2) <sup>6</sup>. |
| Antiviral uptake | 0% | Assuming patients at high-risk of SARS-CoV-2 severe outcomes could be treated timely with antiviral of 90% effectiveness against death with 0% uptake. |
|  | 30% | Assuming patients at high-risk of SARS-CoV-2 severe outcomes could be treated timely with antiviral of 90% effectiveness against death with 30% uptake. |
|  | 70% | Assuming patients at high-risk of SARS-CoV-2 severe outcomes could be treated timely with antiviral of 90% effectiveness against death with 70% uptake. |
| Time scale | Short term | Disease burden after one epidemic wave of SARS-CoV-2 with the epidemic size roughly the same as that of the Hong Kong Omicron wave <sup>7</sup>, i.e., 60% overall infection attack rate. For simplicity, we assume infection attack rates remain the same (60%) across all age groups and vaccination status. We do not consider reinfections. |
|  | Long term | Long term disease burden of a SARS-CoV-2 variant's primary infections, assuming 95% of the population have been infected with SARS-CoV-2. For simplicity, we did not account for the disease burden due to repeated infections after the primary ones. |

**Supplementary Table 3.** The hypothetical origin-destination mobility matrix of NPI intensity Level 0-4.

| Risk level of the origin street/town | Risk level of the destination street/town |  |  | Mobility within one street/town |
| --- | --- | --- | --- | --- |
|  | High | Moderate | Low |  |
| High | 0 | 0.1 | 0.3 | 0.3 |
| Moderate | 0.1 | 0.3 | 0.5 | 0.6 |
| Low | 0.3 | 0.5 | 0.8 | 0.9 |

Note: The risk level refers to the real-time risk level of the street/town at the time of the transmission event occurring.
- 5 The value in each cell of the matrix refers to the average travel probability per person after official report of the first detected infection, given the risk level of the origin and destination region.

**Supplementary Table 4.** The hypothetical origin-destination mobility matrix of NPI intensity Level 5.

| Risk level of the origin street/town | Risk level of the destination street/town |  |  | Mobility within one street/town |
| --- | --- | --- | --- | --- |
|  | High | Moderate | Low |  |
| High | 0 | 0 | 0 | 0 |
| Moderate | 0 | 0.1 | 0.3 | 0.3 |
| Low | 0.1 | 0.3 | 0.5 | 0.6 |

Note: The risk level refers to the real-time risk level of the street/town at the time of the transmission event occurring. The value in each cell of the matrix refers to the average travel probability per person after official report of the first detected infection, given the risk level of the origin and destination region.

